# A Study on the Effects of Containment Policies and Vaccination on the Spread of SARS-CoV-2

**DOI:** 10.1101/2020.10.16.20213835

**Authors:** Vahid S. Bokharaie

## Abstract

This paper presents a method to predict the spread of the SARS-CoV-2 in a population with a known age-structure, and then, to quantify the effects of various containment policies, including those policies that affect each age-group differently. The model itself is a compartmental model in which each compartment is divided into a number of age-groups. The parameters of the model are estimated using an optimisation scheme and some known results from the theory of monotone systems such that the model output agrees with some collected data on the spread of SARS-CoV-2.

To highlight the strengths of this framework, a few case studies are presented in which different populations are subjected to different containment strategies. They include cases in which the containment policies switch between scenarios with different levels of severity. Then a case study on herd immunity due to vaccination is presented. And then it is shown how we can use this framework to optimally distribute a limited number of vaccine units in a given population to maximise their impact and reduce the total number of infectious individuals.

**MSC subclass:** 92C60, 92C50

## 1 Introduction

This manuscript presents a framework to model the spread of SARS-CoV-2 in a population with a known age-structure. The model itself is a compartmental model in which each compartment is divided into a number of age-groups. And a data-driven approach is presented to adapt the parameters of this model to the available data on the spread of SARS-CoV-2. The mathematical framework is an extension of the framework presented in [1] and [2], in which a deterministic compartmental model is used. In such model, the model name represents the progress of the disease under study. For example, in an SIS model, which is the model used in [1] and [2], all individuals initially belong to susceptible compartments (i.e. those who are healthy and can be infected), then Infectious compartment and then when they recover, they join the Susceptible compartment. In other words, it is assumed that there is no immunity to the disease. If the disease includes a latent period (in which the individuals are infected, but not infectious) and if the disease infers immunity, then we should use an SIER model, which is the model more suitable for COVID-19. When I started to work on modelling the progress of SARS-CoV-2 [3], I used an SIR compartmental model. At the time of writing of [3] it was already shown that SARS-CoV-2 infers immunity, for Macaque monkeys in [4] and for humans in [5]. Further investigations by virologists and physicians showed that SARS-CoV-2 also infers an inhibition period, as explained in [6] and references therein. Hence an SEIR model is a more suitable choice than SIR.

Deterministic Compartmental models have been used extensively to model the progress of various diseases, including COVID-19. One recent and notable example is [7], in which an SEIR model is used. Although apart from the general structure of the method, there are not many similarities between their methodology and what is presented in this paper. Fore example, here I have divided each compartment into a number of age-groups, while in [7] no stratification is applied to the compartments. Or as we will see shortly, in the presented method in this paper, the parameter of the age-stratified SEIR model are estimated based on the real world data from a population with a known Basic Reproduction Number, *R*_0_ (defined in Section A.1.2). Subsequently, we can directly apply the effects of various containment strategies to the parameter of the model, independently for each age-group if needed, and then predict the evolution of each compartment in each age-group. But the method in used [7] has used the estimated values of *R*_0_ in United States in a certain time-period to estimate a single contact rate, and then assume it remains constant in the time-period in which they want to predict the evolution of each compartment. An assumption which will be invalid as soon as there is a change in the containment policies in the population.

Apart from the theoretical framework, this work led to two open-source Python libraries. One is called `MiTepid sim` [8] which can be used to simulate the spread of the SARS-CoV-2 in an each age-stratified deterministic compartmental model under any defined containment policy. Another library is called `MiTepid opt` [9] which was used to implement the optimisation scheme presented in this manuscript.

The structure of the paper is as follows. In Section 2, I will explain the outline of the model and the optimisation schemes, without getting into the details. The mathematical details are explained in Appendices A.1 and A.2. Then various case studies are presented in Section 3. They include a study on how changing variables of the model affect the trajectory of the Infectious population. And then studying the effects of various suppression strategies, including those which might affect each age-group differently. Then, long-term containment plans. These are the types of containment policies which we have already seen in many countries around the world. They usually include scenarios in which we switch between policies with different levels of severity. And finally, a study on the effects of vaccination on the population. Using the presented framework, we can see at what level of vaccination we can achieve herd immunity. It will also be shown that if we do not have enough vaccine units to reach herd immunity, how to use the limited number of vaccine units to have a maximum impact. That is another optimisation problem, details of which are explained in Appendix A.3. A discussion on the pros and cons of this approach and possible directions for improving this methodology is presented in Section 4.

## 2 The Method

In the method presented in this paper, an SEIR deterministic compartmental model is used to study and predict the spread of SARS-CoV-2 in any population with a known age-structure. As a reminder, in SEIR models, the population is divided into compartments that capture different stages in the progress of that disease. The four compartments in SEIR model are *Susceptible, S*, which includes those who are healthy and can be infected; *Exposed, E*, those who are infected but not yet Infectious, *Infectious, I*, which includes those who are infected and can transmit the disease; and or *Recovered, R*, also known as *Removed* which includes those who were Infectious but no more. And the term SEIR represents the progress of the disease from S to E to I and to R. If, for example, a virus does not have a latent period but infers immunity, we use SIR compartmental model. There are also some works which have used more detailed compartmental models, for example [10], in which Infectious compartment is further divided into Asymptotic and Symptomatic. In this work, such a differentiation can also be easily incorporated into the model, as will be discussed.

In this manuscript, each compartment is divided into a certain number of groups. The choice of how to group the population is completely arbitrary from a theoretical perspective. But from a practical point of view, such a choice should be informed by the characteristics of the disease and also the availability of the relevant data. And in that regard, a very good choice for SARS-CoV-2 is to divide the population into age-groups. More specifically, the population in each compartment *S, E, I* and *R* is divided into nine age-groups 0-10, 10-20, …, 70-80 and 80+. The subsequent model is a set of 4 *×* 9 nonlinear ordinary differential equations (ODEs) with some unknown parameters which we should estimate based on the known characteristics of the SARS-CoV-2 and the available data. As detailed in Appendix A.1, we can assume the population is constant, which is a reasonable assumptions in the time-scales which are of interest to us. Hence, we end up with a set of 3 *×* 9 ODEs. This set of ODEs have a number of unknown parameters which include *transfer rates, inhibition rates*, and *contact rates*. Transfer rates and inhibition rates, represented with *γ*_*i*_ and *σ*_*i*_ respectively for each of the age-groups *i* = 1, …, 9, can be easily calculated if we know on average, how long is the inhibition period and for how long an individual is Infectious. I have assumed them to be 5 and 4.6 days, respectively, as reported in [6]. And assuming one day to be the unit of time, and assuming this time-period is on average the same for all age-groups, then *γ*_*i*_ = 1*/*5 and *σ*_*i*_ = 1*/*4.6 for *i* = 1, …, 9. Or we can consider different *γ*_*i*_ and *σ*_*i*_ for each age-group if we have such data in each age-group available.

But the real challenge is in estimating the contact rates, represented by *β*_*ij*_ for *i, j* = 1, …, 9. Contact rates denote the rate at which Susceptibles in age-group *i* are infected by Infectious individuals in age-group *j*. Contact rates are very difficult to estimate because they capture various characteristics of the population and the dynamics of the virus. Their value can depend on the average number of direct contacts between the members of different groups, which needs a comprehensive and detailed analysis of the behaviour and mobility of the individuals in the population. Contact rates can also depend on the differences in susceptibility of each age-group to the virus and also on the mechanisms of its transmission. Hence, to directly calculate the values of contact rates is a monumental task that even if possible, might be extremely difficult, time-consuming and expensive.

The main contribution of this work is to show how to overcome this challenge and how to estimate the contact rates. In order to do so, an optimisation scheme is used which is based on two distinct but important sets of data on the spread of COVID-19. One is the estimate for basic reproduction number, *R*_0_, for COVID-19 in an uncontained population. There are various estimates for that parameter, but the one reported in [11], which is *R*_0_ = 2.95 is used in this manuscript. A few other groups have also reported values very close to that [12]. The other piece of information is coming from [13], which shows the relative distribution of confirmed cases of COVID-19 in each age-group in Wuhan, China, as of February 11th, as shown in Table 5 in Appendix A.2. Up until two weeks before this date, Chinese authorities did not impose any meaningful containment strategies and the virus was spreading in an uncontained population. Hence we can assume these numbers are the results of the uncontained spread of the virus.

Using these two sets of information, and using the optimisation scheme as detailed in Appendix A.2, we can estimate the contact rates. One detail of the optimisation scheme which deserves to be highlighted is that it relies on the ratio of the number of reported cases in each age-group, not the absolute values. By doing so, we can avoid two potential issues that might arise in using such data. One is the fact that the number of confirmed cases in each age-group is an unknown fraction of actual Infectious numbers at each time. Another issue is the deliberate or non-deliberate errors that creep in while reporting these numbers. Relying on the ratios of the reported Infected numbers in the optimisation scheme makes it less sensitive to both of these issues, although it does not completely eliminate them. Using more data points as inputs to the optimisation scheme is one possible way to overcome these issues even more.

All the mathematical details of both the model and the optimisation schemes are explained in detail in Appendices A.1 and A.2. The important thing to keep in mind is that although the optimisation scheme uses data collected from China in a certain time-period and in an uncontained population, but the values we obtain can be adapted to any population with a known age-structure and under any containment policy, if we know how that policy affects contacts between various age-groups, or how it will change the *Basic Reproduction Number* (defined in Section A.1.2). The model has some other advantages and also some disadvantages which are discussed in Section 4. Even Considering the disadvantages, it can serve as a good first step to quantify the effects of various containment policies in different countries. The actual values of the contact rates are accompanied with `MiTepid sim` [8], a Python library which can be used to simulate the model used in this paper or any other stratified compartmental model of interest.

## 3 Results

Using the mathematical framework explained in Section 2 and detailed in Appendices A.1 and A.2, we can answer various questions about how SARS-CoV-2 spreads in any population with a known age-structure. We can quantify the effects of various containment policies on the spread of the virus even if those policies affect age-groups differently and if the policies vary in time. It can be used to study the effects of partially vaccinating the population, how wide-spread the vaccination should be to reach herd immunity and how to optimally distribute limited vaccine resources in a population. In this section we will see how we can find answers to all these questions.

### 3.1 Uncontained Spread of the Virus

Let’s consider the case in which SARS-CoV-2 is spread uncontained, i.e. when people in the population interact with each other as in normal times, with no external or self-imposed restrictions in the interactions.

Figures 1a and 1b show the Infectious and Recovered population in each age-group in Germany. As mentioned in the previous section, the output of the model is the evolution of the trajectories of *E*_*i*_, *I*_*i*_ and *R*_*i*_, for *i* = 1, …, *n*. We can then add up these values to obtain the aggregate trajectories of *E, I* and *R* compartments. That is how the Figures 1c and 1d are generated. For each country, the trajectory for each age-group is calculated, and then the aggregate is calculated based on the population distribution for that country. As can be seen, the predicted peak in the number of Infectious people and the eventual ratio of the Recovered population varies considerably among different countries. That can be explained based on differences in the population distribution in these countries. For example, in Iran, 66.8% of the population are under 40 years old while that ratio is 39.8% in Italy [14]. The model predicts that countries with an older population would be affected worse if they let the virus spread uncontained, in agreement with for example [15] when comparing mortality rates in the USA and UK. Table 1 summarises the eventual ratio of Recovered population and the maximum instantaneous Infectious ratio in a few countries.

**Table 1:** Maximum Instantaneous Infectious ratio and eventual Recovered ratio for the spread of SARS-CoV-2 in different countries assuming population is uncontained. The Recovered compartment includes those individuals who have been Infectious and then were recovered or died.

| Country | Eventual Ratio of Recovered Population (%) | Maximum Instantaneous Infectious Ratio (%) |
| --- | --- | --- |
| Germany | 91.77% | 14.35% |
| Iran | 82.15% | 10.33% |
| Italy | 91.23% | 14.21% |
| Spain | 90.35% | 13.95% |
| China | 87.06% | 12.06% |
| USA | 90.56% | 13.63% |
| UK | 91.99% | 14.18% |
| France | 91.95% | 14.32% |
| South Africa | 80.01% | 9.72% |

**Figure 1:**
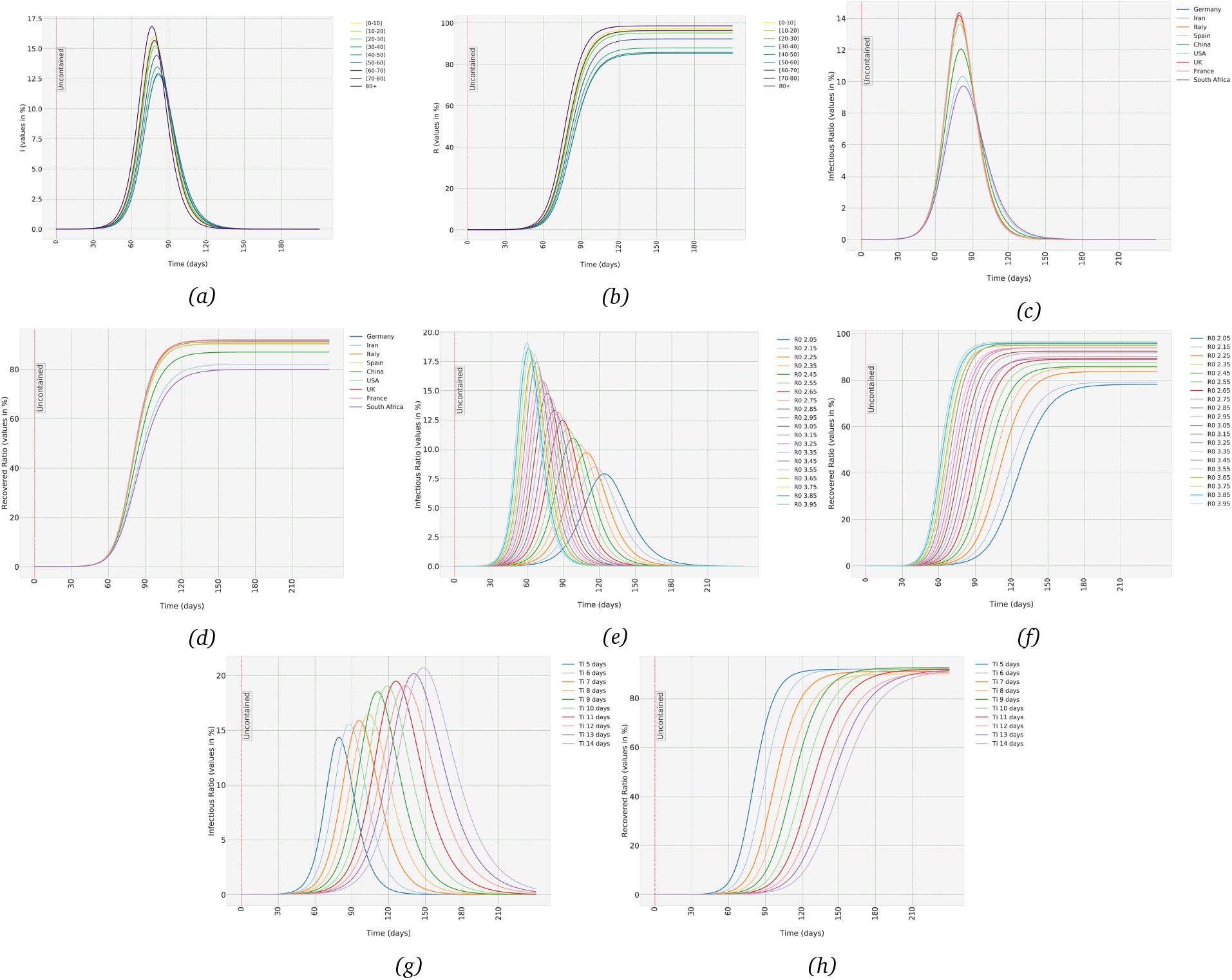
Uncontained scenario: (a,b) The Infectious and Recovered trajectories in the uncontained population in each age-group in Germany, (c,d) Aggregate trajectories of Infectious and Recovered in different countries. (e,f) Aggregate trajectories of Infectious and Recovered in Germany when R_0_ in uncontained population assumes different values, (g,h) Aggregate trajectories of Infectious and Recovered in Germany with different values of T_i_ (the average time-period hosts remain infectious).

To solve any system of Ordinary Differential Equations (ODEs), apart from the equations themselves, we should also define the initial conditions, which in our case means the initial ratios of Infectious, Exposed and Recovered populations in each age-group. In all the figures presented in this manuscript, I have assumed 1 in 100,000 in each age-group is Infectious at time *t* = 0, and the Recovered population is 0. We should be cautious in using a system of continuous ODEs for numbers lower than that. It should be noted that the peak values in Figures 1c and 1d are barely affected by the choice of initial conditions. But that is not true for the time it takes to reach the peak values. Hence, in order to predict the day in which the number of Infectious reaches the peak value, we should have a reasonable estimate of the initial conditions. There have been a few studies so far to estimate the total number of infectious based on the reported confirmed cases or the mortality rates and also the results of seroprevalence studies in different countries. Such studies provide valuable information which allows us to have a reliable estimate of the ratio of Recovered, Exposed and Infectious individuals, and then use these values as initial conditions to calculate the trajectories of each compartment. You can look for example at [16, 17, 18, 19, 20, 21]. Apart from these studies, we can also look at the data collected in the Princess Diamond cruise ship and use it to estimate the number of Infectious individuals in a population based on the number of symptomatic cases. I have reported the summary of Diamond Princess findings in Section A.5 for interested readers.

To test what will happen if *R*_0_ for an uncontained population is different than the value *R*_0_ = 2.95, which is adopted from [11], I have assumed *R*_0_ for uncontained population to be any value in [2.05, 3.95] range with steps of 0.1. We can then estimate contact rates assuming different value of *R*_0_ for an un-contained population and then calculate the trajectories. As can be seen in Figures 1e and 1f, higher *R*_0_ leads to higher peaks in both aggregate Infectious and Recovered trajectories.

Also, I have run the same procedure varying *T*_*I*_, the average time each individual remains Infectious. Figures 1g and 1h show what happens if the actual value of *T*_*I*_ changes in [5, 14] days range. Increasing *T*_*I*_ increases the peak value in Infectious compartment, but delays the time it takes for the Recovered compartment to reach its maximum value.

### 3.2 Suppression Strategies

The advantage of having a stratified model is that we can quantitatively evaluate the effects of various containment strategies that affect different age-groups differently. Table 2 has listed some of the more common policies and how they are defined. The column under *Policy Description* defines to what extent the contacts of age-groups are assumed to change under each policy. These values are chosen intuitively, but any other definitions and any other policies can be easily defined in `MiTepid sim` package which is developed as a part of this study [8].

**Table 2:** Effects of different policies in Germany. Please note that eventual Recovered ratio is the total ratio of the population which have been infected at any time during the spread of the virus.

| Policy Label | Policy Name | Policy Description | Eventual Recovered Ratio (%) | Maximum Instantaneous Infectious Ratio (%) | $R_0$ |
| --- | --- | --- | --- | --- | --- |
| UN | Uncontained | All interactions as in normal circumstances | 91.77% | 14.35% | 2.95 |
| KI | Only Schools and Universities Closed | interactions of [0-20] age-groups decreases to 20% | 81.85% | 11.02% | 2.48 |
| EL | Only Elderly Social Distancing | interactions of 70+ age-groups decreases to 25% | 84.45% | 11.21% | 2.37 |
| KIEL | Schools Closed Elderly Social Distancing | combination of KI and EL | 72.73% | 7.98% | 1.88 |
| KIOF | Schools, Offices and Companies Closed | Combination of KI and interactions of [20-70] age range decreases to 50% | 58.59% | 5.81% | 1.67 |
| ADEL | Adults and Elderly Self-isolate | Interactions of Elderly down to 25% Adults to 20% | 34.19% | 4.29% | 1.05 |
| SD | Social Distancing | combination of KI and EL and interactions of people in [20-70] age range reduce to 20% | 22.27% | 3.94% | 0.63 |
| LD | Lock-down | interactions of all individuals reduced to 10% | 16.41% | 3.84% | 0.30 |

As can be seen in Figures 2a and 2b, all the containment policies decrease both the peak instantaneous Infectious ratio and eventual Recovered ratio, which is to be expected. The most effective policy among the ones listed in the Table 2 is *Lock-down*. It is worth noting that although in Lock-down policy the contact are limited to half of Social Distancing policy, the decrease in the peak Infectious value is much less pronounced. Such observation which are not intuitively predictable, can be valuable for policy-makers to weigh the benefits of a containment policy against its negative impact on the economy and its psychological toll on the individuals in the population.

**Figure 2:**
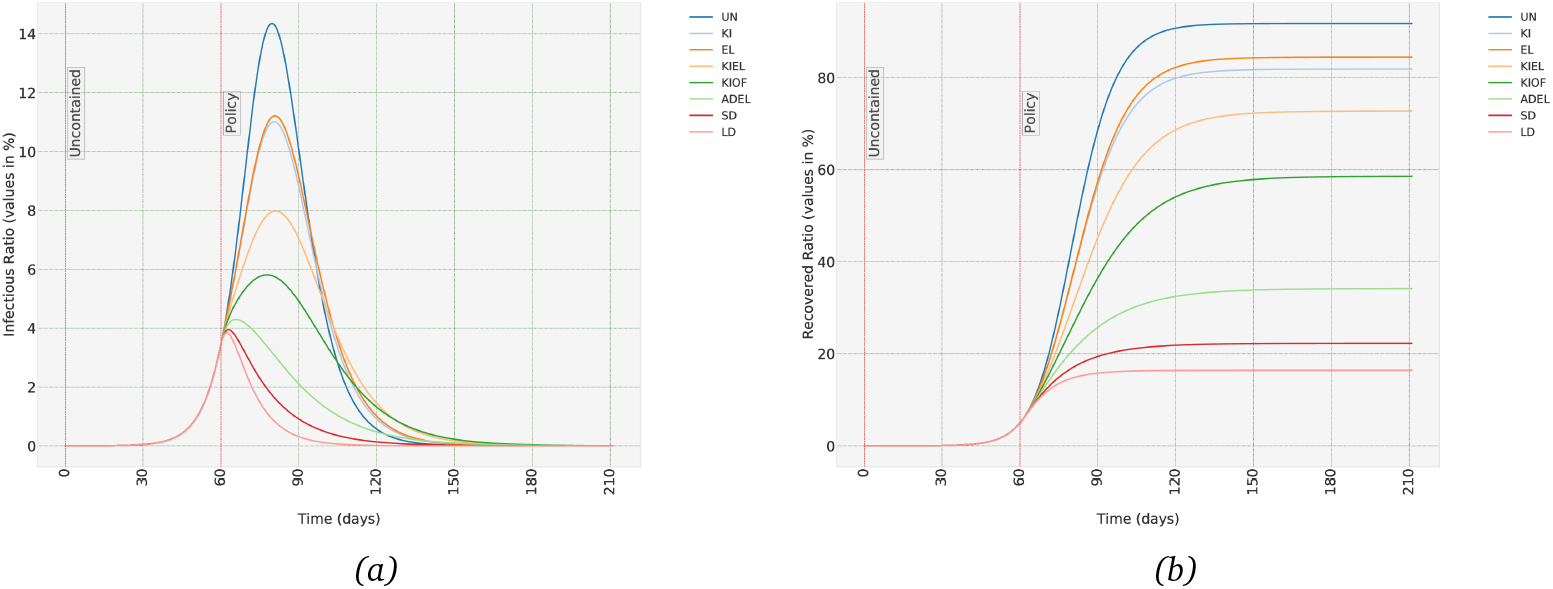
Effects of different suppression policies (as defined in Table 2) in Germany, when population is uncontained for the first 60 days of the spread of the virus. (a) Infectious (b) Recovered trajectory.

Table 2 also shows the basic reproduction number, *R*_0_, for each policies. As can be seen in Figure 2a, and as it is well-known in epidemiology, when *R*_0_ *<* 1, the disease starts to disappear from the population. But as importantly, bringing the ratio of the Infectious population down to a small enough ratio of the total population can take months, even if we impose a total lock-down strategy (the exact time depends on the ratio of Infectious when we start the policy). And maintaining such a strategy might not be feasible in many countries with a more fragile economy. In the next section, we will see what happens if we switch between different strategies with different degrees of severity as a long-term plan.

### 3.3 Mitigation Strategies and Long-term Plans

So far, we have seen what happens if we leave the population uncontained and let the virus spread freely. It can lead to, for example in the case of Germany, around 92% of the population being eventually infected, which can lead to hundreds of thousands of death. Then we saw that some suppression strategies can be hugely effective in containing the spread of the virus and stop such humanitarian disasters. But a complete and long-lasting lock-down strategy, even if possible, can have a huge economic cost, apart from its psychobiological toll on the members of the society. A reasonable compromise is to switch between containment strategies of various degrees of strictness, depending on the observed trend in the number of confirmed cases. What results is what is known in control theory as a *switched system* and Implementing it using this framework is quite easy. We start with the first set of parameters, at time *t* = 0, run the simulations until time *t* = *t*_1_, and use the final state of the previous ODE as the initial conditions for the new one. All of this can be easily implemented in `MiTepid sim` software package [8]. What follows in this section is simply a few examples of how such strategies can or cannot be useful. This by no means is meant to be considered a comprehensive list of effective mitigation strategies but merely examples to highlight the capabilities of this framework. All the examples discussed in the following are based on the population distribution of Germany.

Let’s start with a case in which we switch between uncontained case and Social-Distancing, as defined in Table 2. It means the case in which contacts between children and adults is brought down to 20% of uncontained case, and for elderly to 25%. Since for uncontained we have *R*_0_ = 2.95, in Social Distancing it decreases to *R*_0_ = 0.63. We assume uncontained case for the first 30 days, starting from the initial state in which 1 in 100,000 is Infectious. As can be seen in Figures 3a and 3b the policy manages to initially contain the virus, but as the number of Infectious during uncontained phases increase, the exponential growth causes the number of Infectious to reach its peak value at 150 days and then start to decrease. Although this policy is obviously not able to contain the number of Infectious in a manageable level, we can still see some benefits in it. The peak value of Infectious is now less than what it was in uncontained case, 9.6% vs 14.3%. Also, the time to reach the peak value has increased from around 80 days to 150 days. But more importantly, the eventual recovered ratio, i.e. the total number of individuals who were infected at some stage, is decreased from around 92% to around 62%. That means although this policy will eventually lead to, most probably, overload of the health system resources, but it can limit the total death toll significantly.

**Figure 3:**
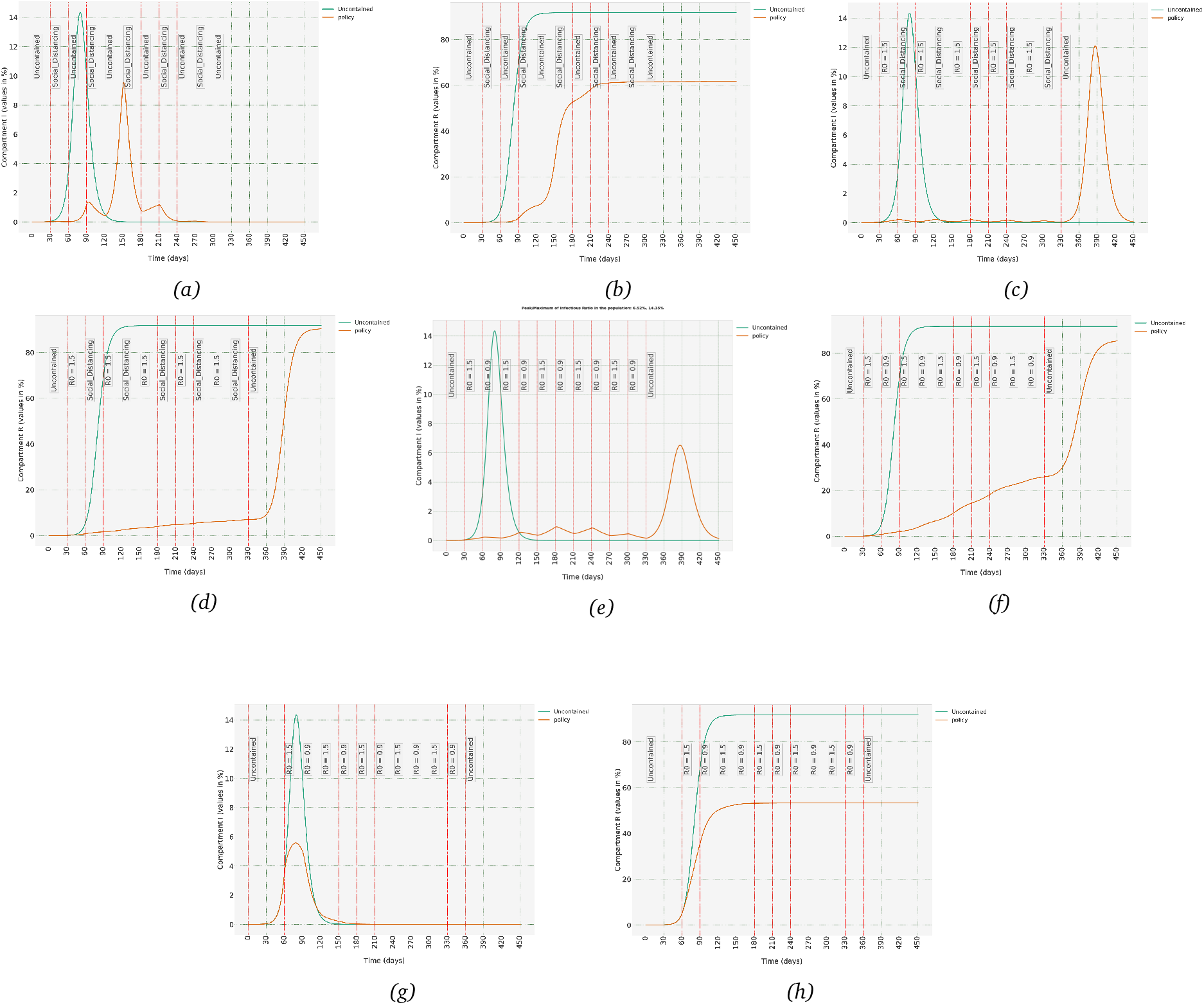
The effects of long-term strategies on Infectious and Recovered trajectories when we switch between policies of different degrees of severity. Polices as defined in Table 2: (a,b) Switching between uncontained and social-distancing scenarios, (c,d) A less strict long-term strategy when we switch between social distancing and R_0_ = 1.5 scenarios, (e,f) switch between situations where R_0_ = 0.9 and R_0_ = 1.5. Containment policy can contain the spread of the virus when imposed 30 days after 1 in 100,000 in population is Infectious, (g,h) Same as in Figure 3e, only difference is that policy is imposed 60 days after Infectious population reaches 1 in 100,000. An extra 30 days of inaction causes the ratio of Infectious to reach a peak of 5.58%.

Now let’s repeat the same scenario, but now, after the first 30 days of uncontained policy, we replace uncontained policy with the one in which *R*_0_ = 1.5. Given *R*_0_ = 2.95 for uncontained population, this means contacts between members of the population is assumed to be cut to almost half. Figures 3c and 3d show how Infectious and Recovered populations changes. This time, the policy is successful in containing the spread of the virus. Obviously, it does not make sense to go back to uncontained at 330 days, but I have left it there so we can see the potential the population has for a new outbreak if we relax the restrictions.

But reaching the same level of restrictions as we have defined under the Social-distancing policy can be very difficult and costly. So, let us look at a more realistic scenario, in which after the first 30 days of uncontained population, we switch between cases in which *R*_0_ = 1.50 and *R*_0_ = 0.9, which is not far from what has already happened in some countries. Figures 3e and 3f show the outcome. It is no surprise to see that the number of Infectious is now more than the previous case but still the maximum number of Infectious is far from that of the uncontained scenario. But what happens if we start imposing this switching policy after 60 days of uncontained population, not 30 days. Figures 3g and 3h shows the price we have to pay for an extra 30 days of inaction in containing the spread of the virus. Ratio of Infectious starts to get out of hand, and although the peak is much less than uncontained case 5.58% vs 14.35%, it can still be overwhelming for public health resources.

There are various other possibilities that can be studied using this framework, all of which can be easily implemented using the `MiTepid sim` Python package [8].

### 3.4 Herd Immunity due to Vaccination

In Section 3.1 we saw that even in the uncontained case, not all of the individuals in the population need to be infected for the spread of the virus to be stopped. As can be seen in Table 1 and Figure 1d, in an uncontained population in which SARS-CoV-2 is spread freely, when the total number of recovered reaches a certain value, the disease dies out, a phenomenon which is called *Natural Herd Immunity*. The exact value depends on the dynamics of the virus and also age-structure of the population in case of SARS-CoV-2. But the price we pay for natural herd immunity is catastrophic. For example, in Germany, natural herd immunity is reached when around 92% of the population is eventually infected, a parameter that I call *Herd Immunity Threshold (HIT)*. And with a mortality rate of 0.6% that amounts to around 460 thousand deaths.

We have already seen in the previous section that imposing any kind of containment policy, can reduce the HIT significantly. Now let us see how introducing a vaccine can changes the HIT. In particular, we want to know what ratio of the population should be vaccinated to reach herd immunity? Answering that question is very easy using the framework we have in place. There is no need to define a new compartment for the vaccinated people. We can assume the vaccinated population as the initial conditions for the Recovered compartment and then see how the trajectories of Infectious and Recovered evolve.

Table 3 shows the eventual Recovered and maximum Infectious ratios under different initial ratio of vaccinated population, assuming that after vaccination, a containment strategy is in place such that *R*_0_ = 1.50. Remember that *R*_0_ = 2.95 in an uncontained population, hence *R*_0_ = 1.50 means interactions in the populations are reduced to around half of that in an uncontained case. As can be seen in Table 3, when we inoculate around 30% of the population, we have already reached the herd immunity level. If we leave the population uncontained, we need to vaccinate around 65% of the population to reach HIT. Please note that these values are for a population with the same age-structure as Germany, for countries with a younger population, HIT is lower.

**Table 3:**
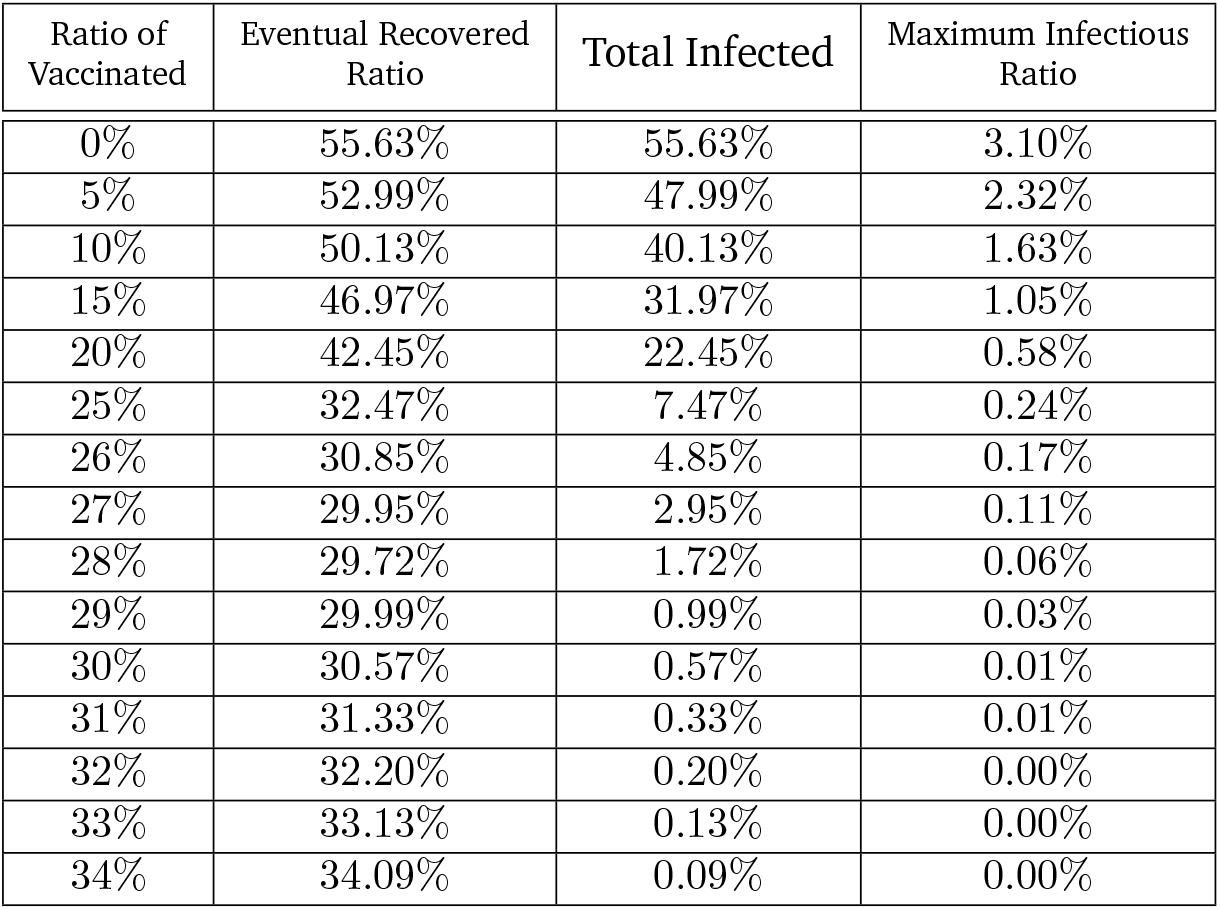
How vaccinating different ratios of the population of Germany changes the total and maximum Infectious Ratios. It is assumed the population is under a containment policy that keeps R_0_ = 1.50.

| Ratio of Vaccinated | Eventual Recovered Ratio | Total Infected | Maximum Infectious Ratio |
| --- | --- | --- | --- |
| 0% | 55.63% | 55.63% | 3.10% |
| 5% | 52.99% | 47.99% | 2.32% |
| 10% | 50.13% | 40.13% | 1.63% |
| 15% | 46.97% | 31.97% | 1.05% |
| 20% | 42.45% | 22.45% | 0.58% |
| 25% | 32.47% | 7.47% | 0.24% |
| 26% | 30.85% | 4.85% | 0.17% |
| 27% | 29.95% | 2.95% | 0.11% |
| 28% | 29.72% | 1.72% | 0.06% |
| 29% | 29.99% | 0.99% | 0.03% |
| 30% | 30.57% | 0.57% | 0.01% |
| 31% | 31.33% | 0.33% | 0.01% |
| 32% | 32.20% | 0.20% | 0.00% |
| 33% | 33.13% | 0.13% | 0.00% |
| 34% | 34.09% | 0.09% | 0.00% |

Figures 4a and 4b show the evolution of Infectious and Recovered trajectories when we start from different ratios of vaccinated population when we keep *R*_0_ = 1.5 in our population and Figures 4c and 4d show evolution of trajectories for an uncontained population. But in these figures, we have assumed the same ratio in each age-group is vaccinated. In other words we have ignored an important capability of the model, the ability to impose different conditions on different age-groups. So, let’s say we have enough vaccine units to only inoculate 15% of the population of Germany. We can turn this problem into an optimisation problem, in which we keep overall number of vaccinated population at 15% while vaccinating different ratios of each age-group with the aim of minimising the eventual Recovered population. Table 4 shows the difference that non-uniform vaccination in age-groups can have and Figures 4e and 4f show the evolution of trajectories corresponding to uniform and optimised cases. Obviously, it is not really realistic that we vaccinate only 15% of the population and then none. This simple example is merely used to highlight this feature of this framework. The code used to run this optimisation approach is also available in `MiTepid opt` [9] package. Other cases for different populations and different and even time-varying numbers of avilable vaccine units can be studied using `MiTepid opt`.

**Table 4:** Difference between the case in which 15% of each age-group in the population of Germany is vaccinated and the optimised case, in which different fractions of each age-group are vaccinated, but 15% of the population in total. Population is assumed to be under a containment policy that keeps R_0_ = 1.50.

| case | Eventual Recovered | Total Infected | Maximum Infectious |
| --- | --- | --- | --- |
| uniform | 46.97% | 31.97% | 1.05% |
| optimised | 44.28% | 29.28% | 0.87% |

**Figure 4:**
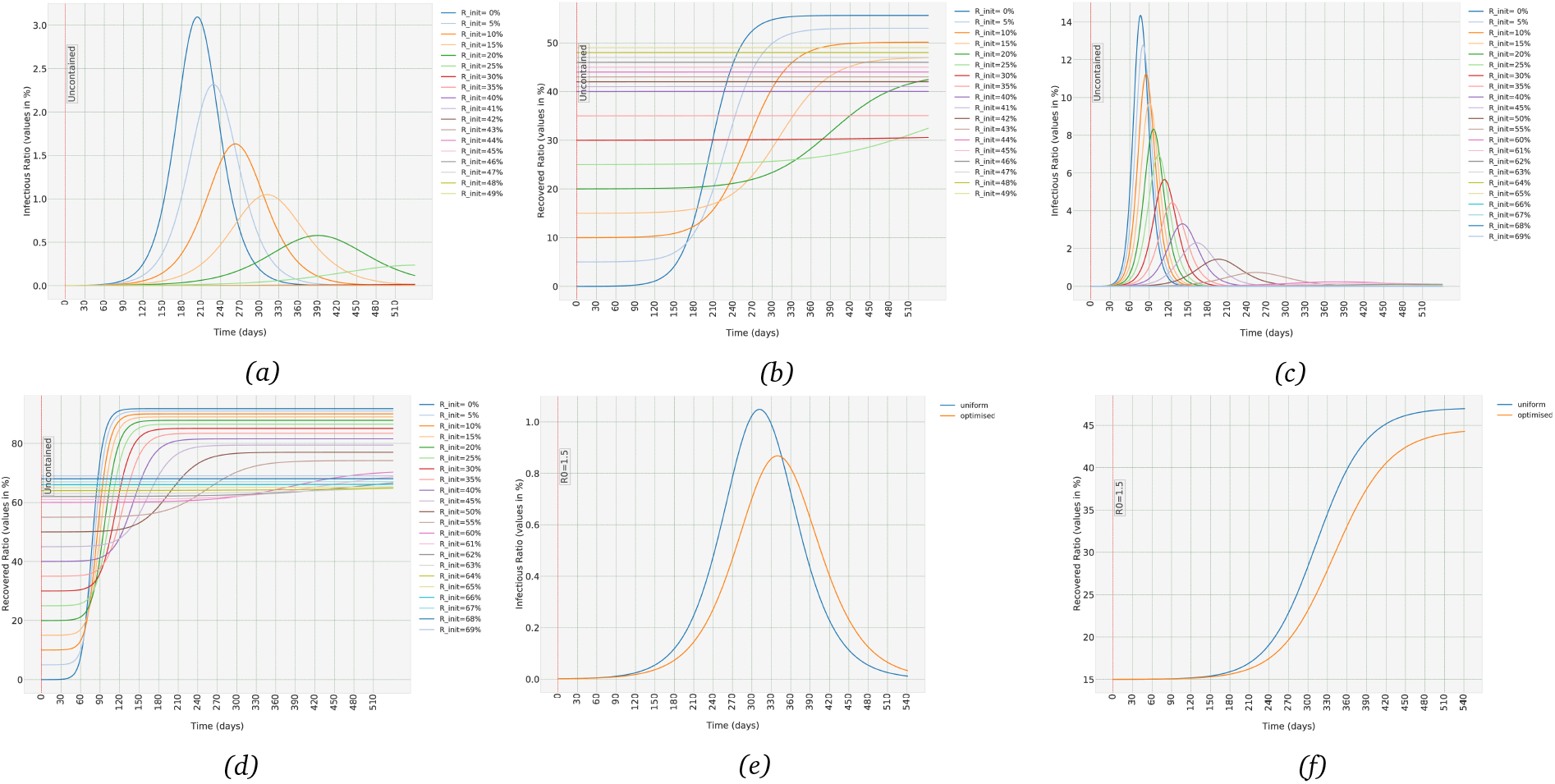
(a,b) The effect of vaccinating different ratio of the population of Germany on the evolution of the Infectious and Recovered trajectories. It is assumed population is contained to R_0_ = 1.5, (c,d) Same as (a) and (b), but it is assumed the population is uncontained, (e,f) Difference between uniform vaccinations and optimally distributed vaccination in Infectious and Recovered ratio when we have enough vaccines to vaccinate only 15% of the population. Optimal distribution of the vaccines allows us to end up with around 2.7% reduction in the total number of infected.

## 4 Discussions

The method presented in this manuscript can be used to predict the spread of SARS-CoV-2 in each age-group of a population with a known age-structure. Not just the aggregate values, but also the evolution of the number of Susceptible, Infectious, Exposed and Recovered individuals in each age-group. It was shown how we can use various containment strategies and how to quantify their effects on the evolution of the trajectories. We saw that the method can incorporate strategies which might be time-varying and might affect each age-group independently. The main advantage of this framework is that it allows us to estimate the contact rates of the model without a detailed and explicit knowledge of how and to what extent different age-groups in a population interact with each other, or how the virus affects each age-group. The contact rates were estimated based on the available data on the spread of COVID-19. The model itself is a set of nonlinear Ordinary Differential Equations (ODEs), hence simulating various containment policies in different time frames does need any special computational power, which makes this methodology attractive to researchers or policy-makers who do not access to high performance computing facilities. Also, it should be noted that we can stratify the compartments not necessarily to age-groups, but based on any other stratification of interest.

To better estimate the contact rates, any other available data can be used as the input to the optimisation scheme. In this methodology, to estimate the contact rates in any population, we need only two points in time with a known number of reported infected cases in each age-groups while the Basic Reproduction Number, *R*_0_, remains constant and known in between the two time-points. As explained in Section A.2, we can add more of such time-points to make the estimates more accurate. As already mentioned, one major advantage of this methodology is that if we obtain such estimation from data collected in China, which is the case for the estimates presented in this paper, we can easily adapt them to any other country/region/city with a known age-structure.

In spite of all its strength, the methodology has some shortcomings that should be kept in mind while interpreting the results. The method assumes the virus affects individuals (of the same age) in different countries similarly. If it is the case that the differences in genetic backgrounds or vaccination histories in different countries can affect the infection rate, such differences would be lost while transforming the contact rates from one population to another. Also, it is an implicit assumption in the model that the general interactions between people in different countries and societies are in general similar to each other. To clarify the point, if, for example, people older than 70 years in one country live with or have more contacts with the younger age-groups while in another country they live in isolation or in nursing homes, then differences in contact rates caused by such social and cultural differences would be lost. But we can turn this disadvantage into a tool to discover such cultural differences in different populations. If we estimate the contact rates based on two separate set of data collected in two different countries, and then compare the resulting normalised contact rates, we can discover such minute differences in how different age-groups in the two populations interact with each other.

Lastly, it is worth discussing some of the issues which have proven to be important during the SARS-CoV-2 pandemic, and their relationship to the presented methodology. One is the issue of Asymptomatic/Symptomatic infected individuals and how that can be incorporated into this model. Regardless of how and to what extent asymptomatic infected individuals can infect susceptible individuals, and how severity of symptom can affect that, we can easily incorporate such distinctions between individuals in the Infectious compartment into the model. The solution is to simply split the Infectious compartment into two subsets, *I*_*A*_ and *I*_*S*_ for Asymptomatic and Symptomatic. And we should have an estimate of what percentage of each age-group belong to *I*_*A*_ or *I*_*S*_, which as we saw earlier, could be done, for example, using the data collected from Princess Diamond cruise ship as explained in Section A.5. In such a case, we run the same optimisation scheme but number of parameters to be estimated increases, which merely add to computational load of the optimisation scheme. The other issue is mass-testing in a population. Seroprevalence tests can provide valuable information and allows us to more accurately estimate the contact rates. Wide-spread seroprevalence tests at any give time can give us a reliable estimate of the value of Recovered and Infectious compartment at that time which we can then use as extra time-points in our optimisation scheme, as detailed in Section A.2. The other issue is contact tracing, which has also been used in many countries. The premise of contact-tracing is to find the direct contacts of an infected individual and then ask them to self-isolate. This subset of Infectious group can be defined as a new compartment, and then we can treat this new compartment the same way we discussed *I*_*A*_ and *I*_*S*_ compartments.

The method can be extended in various directions to make the results even more useful. We can, for example, divide each age-group into sub-groups based on their vulnerability to the virus, or based on the relative amount of interactions with other individuals in the population. An obvious choice is people who work from home or are unemployed and people who have work at offices. Or people in 60-69 age range who are retired versus those who still work. Even a rough estimate of the relative numbers of these sub-groups in each age-group can give us more insight into more effective ways to contain the spread of the virus with less social and economic impact.

Given the fact that nowadays the population structure in different cities/regions in almost every country is known, we can use the model to describe the spread of the virus for each region or city, and then, assuming in and out-flow traffic to each city is known, consider them as exogenous inputs to our switched system. By doing so, we can have a more detailed and accurate estimate of the spread of the virus in wider geographical areas. That would allow us to consider time-lags that might exist in the spread of COVID-19 in different parts of a country. But even as it is, this model and its estimated parameters can be a useful tool for different policy-makers in different countries in particular those countries with limited computational resources or not enough people with expertise in mathematical epidemiology.

## Data Availability

All data can be found in github.com/vahid-sb/MiTepid_sim

## Declarations

### Ethics approval

N/A

### Consent to participate

N/A

### Consent for publication

N/A

### Availability of data and material

N/A

### Code availability

The code used to simulate the model and prepare the figures is publicly available in: https://github.com/vahid-sb/MiTepid_sim. The code used for optimisation schemes is publicly available in: https://github.com/vahid-sb/MiTepid_opt.

### Authors’ contributions

V.S.B. designed the study, implemented it and prepared the manuscript.

## Appendices

### A.1 The Mathematical Model

To understand the model discussed in this section, it is enough to know some basic concepts in Ordinary Differential Equations (ODEs) and linear algebra. Knowing the following notations and definitions can be helpful.

#### A.1.1 Notations and Some Basic Definitions

ℝ is the field of real numbers and ℝ_+_ is The set of non-negative real numbers. ℝ^*n*^ is The space of column vectors of size *n* of real numbers and ℝ^*n×n*^ is The space of *n × n* matrices of real numbers. I use *x*_*i*_ to represent The *i*th entry of the vector *x* in ℝ^*n*^, for *i ∈{*1, *…, n}*. Please note that *x*_0_ is a vector in ℝ^*n*^ that usually represents initial condition. Notation *a*_*ij*_ is used for (*i, j*) entry of the matrix *A. D* = diag (*x*) is an *n × n* diagonal matrix in which *d*_*ii*_ = *x*_*i*_ for all *i. A*^−1^ is The inverse of the matrix *A. I* is the identity matrix of proper dimensions and 0 is the zero matrix of proper dimensions. *σ*(*A*) is the set of all eigenvalues (spectrum) of the matrix *A. ρ*(*A*) is the spectral radius of the matrix *A*, i.e. the maximum of the absolute values of all eigenvalues.

*A ≫ B* means *a*_*ij*_ *> b*_*ij*_, for all *i, j ∈ {*1, …, *n}*. It should not be mistaken with Positive Definite (PD) matrices. *A > B* means *a*_*ij*_ *≥ b*_*ij*_, for all *i, j ∈ {*1, …, *n}* and *A* = *B* and *A ≥ B* means *a*_*ij*_ *≥ b*_*ij*_, for all *i, j ∈ {*1, …, *n}*. 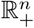 is The positive orthant of ℝ^*n*^, given by *{x ∈* ℝ^*n*^ : *x ≥* 0*}*.

A matrix *A* is called *Hurwitz*, if *µ*(*A*) *<* 0. A real *n × n* matrix *A* = (*a*_*ij*_) is *Metzler* if its off-diagonal entries are non-negative.

The matrix *A* is *irreducible* if and only if for every non-empty proper subset *K* of *N* := *{*1, …, *n}*, there exists an *i ∈ K, j ∈ N \ K* such that *a*_*ij*_ = 0. When *A* is not irreducible, it is *reducible*.

For any subset 𝒰 of ℝ^*n*^, a point *x*_0_ is called an *interior point* of 𝒰 if there is an open ball around *x*_0_ which is wholly contained in 𝒰. The set of all interior points of 𝒰 is called the interior of 𝒰 and is denoted by int (𝒰).

#### A.1.2 SIS Model

The SIS model, although not used in the main text, is presented here, both in the interest of completeness and to provide a theoretical basis for the SIR/SEIR discussions. The formulation presented in this section is adopted from [1] and [2]. In SIS model, the population of interest is first divided into two compartments S, Susceptibles, and I, Infectious. Each compartment can be sub-divided into *n* groups. These groups can represent different age-groups, different health conditions, professions, etc. In this manuscript, I have divided the population in each compartment into *n* = 9 age-groups defined as 0-10, 10-20, …, 70-80 and 80+.

Let *I*_*i*_(*t*) and *S*_*i*_(*t*) be the number of Infectious and Susceptibles at time *t* in group *i* for *i* = 1, *…, n*, respectively. Also, let *N*_*i*_(*t*) = *S*_*i*_(*t*)+ *I*_*i*_(*t*) be the total population of group *i*. The total population of each group is assumed to be constant; formally, *N*_*i*_(*t*) = *N*_*i*_. This does not oversimplify the model, especially when the total population is significantly greater than the number of dead and newborn. But even if that assumption is not deemed realistic for a population, the formulation stated below can still be used as we will shortly see.

Here, *β*_*ij*_, the contact rate between groups *i* and *j*, denotes the rate at which Susceptibles in group *i* are infected by Infectious in group *j* for *i, j* = 1, *…, n*. Further, *γ*_*i*_, the transfer rate, is the rate at which an infectious individual in the group *i* leaves the Infectious compartment and join the Susceptible compartment. We also consider birth and death in the population, but as discussed, we set the birth and death rates in each age-group to be the same value *µ*_*i*_ to keep the total population in each group constant. Using the mass-action law, the basic SIS model is then described as follows [1]:

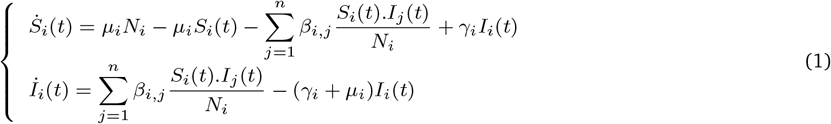

Since the population of each group is constant, it is sufficient to know *I*_*i*_(*t*). If we set *x*_*i*_(*t*) = *I*_*i*_(*t*)*/N*_*i*_ and 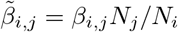, we obtain the following set of differential equations:

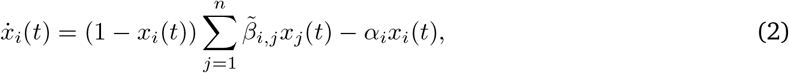

for all *i* = 1, …, *n*. By definition, *x ∈ B*_*n*_ where 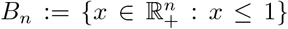. We can write the set of differential equations (2) in compact form as:

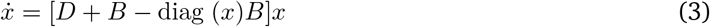

where *D* = − diag (*α*_*i*_) and 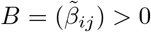.

The following properties of (3) are easy to check. Interested reader can look at [2] for proofs.

i. *f* (*x*) = [*D* + *B* − diag (*x*)*B*]*x* with *D* and *B* defined as above is *C*^1^ in ℝ^*n*^, therefore, the solution for every initial condition in ℝ^*n*^ exists and is unique for all *t ≥* 0.
ii. The origin is an equilibrium point of (3). This equilibrium is referred to as the disease-free equilibrium of the system (3).
iii. System (3) may have an equilibrium in int 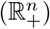 (also referred to as an endemic equilibrium). Conditions for existence of endemic equilibrium for the system (3) depends on parameter *R*_0_, defined below.

One important parameter in mathematical epidemiologically is the *basic reproduction number, R*_0_, which is defined as follows.

##### Definition A.1.1 (Basic reproduction number)

*The basic reproduction number is the expected number of secondary cases produced, in a completely susceptible population, by a typical infective individual during its entire period of Infectiousness [22]*.

For the SIS model (3), following [1], it can be proved that *R*_0_ = *ρ*(*D*^−1^*B*). The reproduction number can be used to characterise the existence and stability of the equilibria of (3). As shown in [1, Theorem 2.3], the disease-free equilibrium, i.e. the origin, is a globally asymptotically stable equilibrium of the system (3) if and only if *R*_0_ *<* 1 (if matrix *B* is irreducible). And the endemic equilibrium, an equilibrium in int 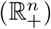, is globally asymptotically stable if and only if *R*_0_ *>* 1. In other words, the necessary and sufficient condition to eradicate a disease for a population in an SIS model is to satisfy *R*_0_ *<* 1.

#### A.1.3 SIR Model

The SIR model is quite similar to SIS, with a minor difference, namely, those who are cured, join the Recovered, *R*, population, not *S*. Hence, the formulation for an SIR model is as follows:

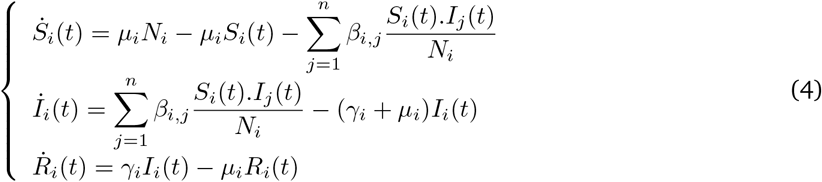

Again, assuming *N*_*i*_(*t*) = *S*_*i*_(*t*) + *I*_*i*_(*t*) + *R*_*i*_(*t*) is constant, similar to what was done in the previous section, if we set *x*_*i*_(*t*) = *I*_*i*_(*t*)*/N*_*i*_ and *y*_*i*_(*t*) = *R*_*i*_(*t*)*/N*_*i*_ and 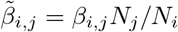 and *α*_*i*_ = *γ*_*i*_ + *µ*_*i*_, we obtain the following differential equation:

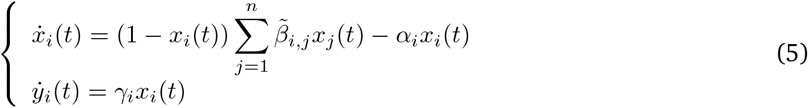

*∀i* = 1, …, *n*. In compact from, (5) can be written as follows:

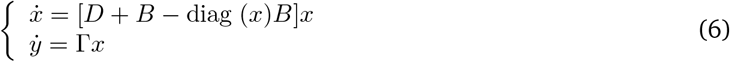

where *D* = −diag (*α*_*i*_) and 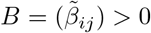 and Γ = diag (*γ*_*i*_) for *i* = 1, …, *n*.

The system (6) has the following properties.

i. *f* (*x*) = [*D* + *B* − diag (*x*)*B*]*x* and *g*(*y*) = Γ*x* with *D, B* and Γ defined as above are *C*^1^ in ℝ^*n*^, therefore, the solution for every initial condition in ℝ^*n*^ exists and is unique for all *t ≥* 0.
ii. To calculate the equilibria of the system we set *f* (*x*) = 0 and *g*(*x*) = 0. One equilibrium is the origin, Disease-Free equilibrium, and the other one, corresponds to the case in which the disease has swept through the population and a significant ratio of the population (and not necessarily all the population) belongs to Recovered compartment and I=0.
iii. Basic Reproduction Number for (6), can be calculated using the same formula *R*_0_ = *ρ*(−*D*^−1^*B*), when all *R*_*i*_ and *I*_*i*_ are close to 0.

Property (iii) follows from the discussion in [23, Section 3] and the fact that this equation is derived from the model linearised around the origin. This also means that as the Infectious ratio increases, the effective *R*_0_ becomes less than *ρ*(−*D*^−1^*B*).

#### A.1.4 SEIR Model

The SEIR model is an extension of SIR in which the susceptibles enter a Exposed, *E*, compartment, before becoming Infectious. The formulation of an SEIR model can be stated as follows:

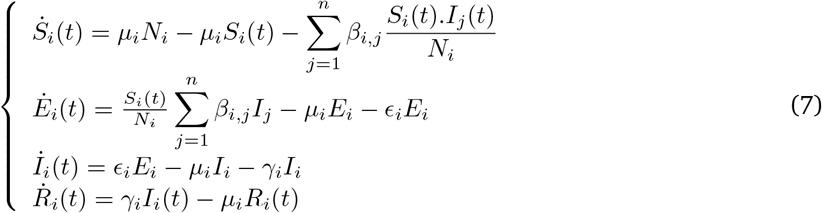

Again, we assume *N*_*i*_(*t*) = *S*_*i*_(*t*) + *E*_*i*_(*t*) + *I*_*i*_(*t*) + *R*_*i*_(*t*) is constant. Now we define *x*_*i*_(*t*) = *I*_*i*_(*t*)*/N*_*i*_ and *y*_*i*_(*t*) = *R*_*i*_(*t*)*/N*_*i*_ and *z*_*i*_ = *E*_*i*_*/N*_*i*_ and 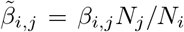 and *α*_*i*_ = *γ*_*i*_ + *µ*_*i*_, we obtain the following differential equation:

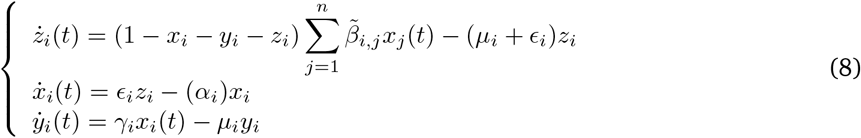

*∀i* = 1, …, *n*. In compact from, (8) can be written as follows:

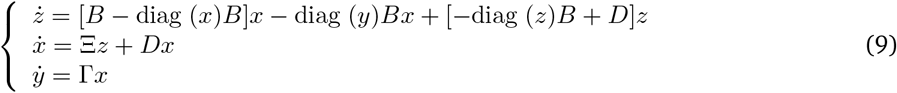

where *D* = −diag (*α*_*i*_) and 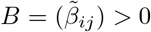 and Γ = diag (*γ*_*i*_) and Ξ = diag (_*i*_) for *i* = 1, *…, n*.

The same as in 6, the origin is also an equilibrium for 9. And using the same arguments as presented in the previous Section, we can use *R*_0_ = *ρ*(−*D*^−1^*B*) as a reasonable estimate for Basic Reproduction Number, *R*_0_ when the trajectory is around the origin. When the trajectory moves away from the origin, *ρ*(−*D*^−1^*B*) over-estimates the actual value of *R*_0_.

### A.2 An Optimisation Scheme to Estimate the Parameters of the Epidemiological Model

This section explains the optimisation method used to estimate the contact rates for an age-stratified compartmental model during the spread of SARS-CoV-2. In order to solve ordinary differential equations (3), (6) or (9), we need to have a reliable estimate of 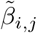 and *γ*_*i*_ for all *i, j*. Parameters *γ*_*i*_ and *ϵ*_*i*_ are easy to estimate. If for example the average duration that individuals in a group *i* are Infectious is 5 days, then *γ*_*i*_ = 1*/*5 = 0.20, given that we have chosen one day to be the unit of time. Estimating 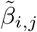, on the other hand, is very difficult, and this section explains how we can estimate contact rates based on real-world data on the spread of COVID-19.

The optimisation method uses some known properties of ordinary differential equations and also results from linear algebra. Let’s say we use an SEIR compartmental model and divide our population into 9 age-groups. Hence, we have 81 values of 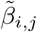 which should be estimated. Let’s say we have two points in time, *t*_0_ and *t*_*f*_ in which we know the ratio of infectious in each age-group. In other words, we know *x*(*t*_0_) and *x*(*t*_*f*_), but only using these two time-points, we end up with infinitely many estimates for contact rates. We need extra information to limit the range of estimated values. One possibility is to add more time-points in which we have a reliable estimate of total number of infectious. The other possibility, in case the containment policy applied in between time-points *t*_0_ and *t*_*f*_ was unchanged and Basic Reproduction Number known, is to use the equations *R*_0_ = *ρ*(*D*^−1^*B*), which we obtained in the previous section. Hence, the extra constraint in the optimisation scheme is that matrix *B* should be chosen such that *ρ*(− *D*^−1^*B*) is equal to estimated value of *R*_0_ in that population.

In this paper, I have used the data collected from Wuhan province in China on February 11th, 2020 [13]. During this time-period, the Basic Reproduction Number, *R*_0_, was estimated to be *R*_0_ = 2.95. For the initial conditions, based on the report that the virus has started to spread in the population in late November, I assumed that 75 days before February 11th, 1 in 100,000 in the population has been infected. Having these information, we can use any global optimisation scheme to estimate the values of the matrix *B* in (9). One possible issue with this approach is that it can be quite sensitive to the reported values at time *t*_*f*_. Such inaccuracies, caused by deliberate or non-deliberate errors in reporting the number of infectious, are quite common. To overcome this issue, I slightly altered the optimisation scheme: instead of trying to estimate the contact rates such that values of *x*(*t*_*f*_) match the real data, we can estimate them such that the ratio of the elements of vector *x*(*t*_*f*_) match that of the real data. Table 5 is used for that purpose. Column C2 in Table 5 shows the distribution of confirmed cases of COVID-19 in different age-groups in Wuhan as of Feb. 11th [13]. We should then normalised these numbers to the relative distribution of each age-group in the general population, the same way that we have normalised each variable in (8) to the total population in each age-group. That is equivalent to dividing each row in Column C2 to corresponding row in column C1, which leads to values we can see in second last column of the Table 5. The last column shows the exact same ratios, normalised to the smallest value in the column, which happens to be the first row. Hence, the optimisation scheme as used in this paper can be summarised as follows, assuming we use the SEIR model.

**Table 5:**
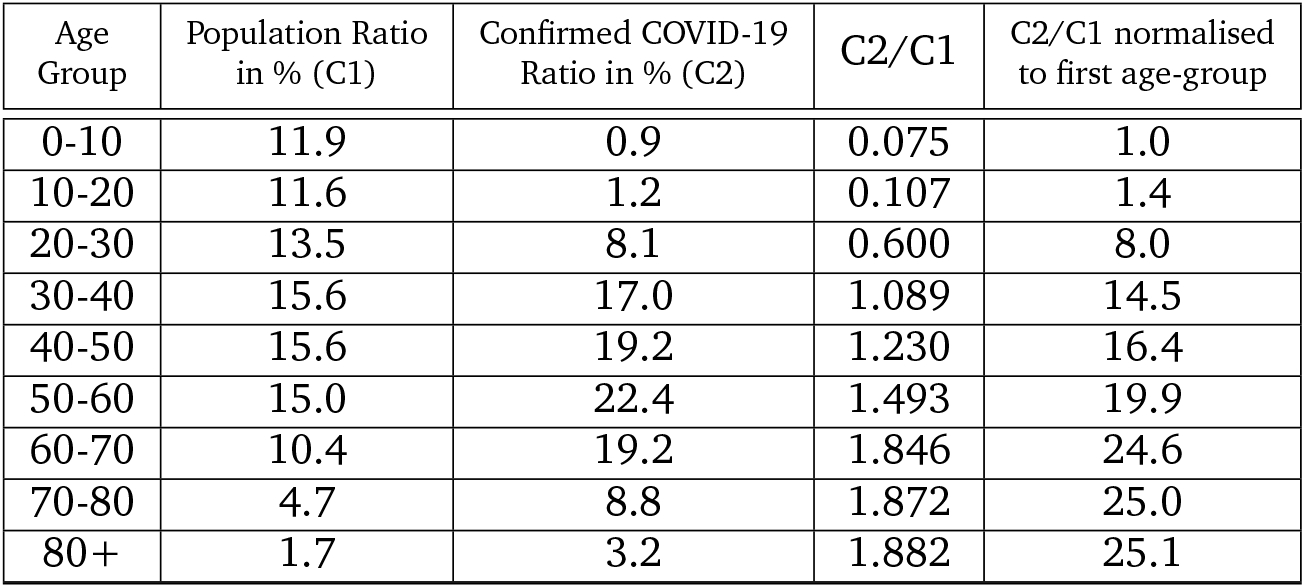
Population distribution in China and the distribution of Confirmed COVID-19 cases in China as of Feb. 11th. To normalise the distribution of confirmed cases, we can divide the values over the population ratio of that age-group. The resulting values are then used in the optimisation scheme designed to estimates the parameters of the epidemiological models.

Find the matrix *B* in (9), such that:

i. For a given dialogical matrix *D* and scalar *R*_0_ = 2.95, *R*_0_ = *ρ*(−*D*^−1^*B*)
ii. The relative values of the states of system (9) at time *t*_*f*_ with initial condition *x*_0_ satisfy the values of the last column of Table 5.

Now that all the required parameters are set, we can solve the optimisation problem to find a *B*_*opt*_. In order to solve the optimisation problem, I have used `sqp` algorithm in `globalsearch` function in `Global Optimisation Toolbox` in Matlab^©^. Optimisation is done in two steps, in the first step, initial values for matrix *B* are chosen randomly from a uniform distribution. When the optimisation algorithm converges to a solution, the optimisation procedure is repeated, this time with the optimum values obtained in the first step as the initial values.

The objective function in the optimisation scheme is the weighted sum of two terms. One is the 2-norm of the difference between the ratio of trajectories of the set of ODEs at time *t*_*f*_ and the desired ratios extracted from Table 5. The second term is the difference between *ρ*(−*D*^−1^*B*) and the desired basic reproduction number of *R*_0_ = 2.95. The weights are tuned with trial and error such that none of the terms is obscured by the other one during the optimisation scheme. Also, the feasible values for the elements of the matrix *B* is set to be in [0.01, 0.20] range. Without such an explicit constraint, the deviation in the values of the optimised contact rates can be unreasonably high, which is a side-effect of using the global optimisation method.

To implements the optimisation scheme, I have used an in-house open-source software called `MiTepid opt` [9]. The optimisation algorithm runs in 15-20 minutes on 40 hyper-threaded CPUs of type Intel(R) Xeon(R) CPU E5-2687W v4 @ 3.00GHz.

**Note A.2.1** *It should be noted that the values obtained from the optimisation schemes are* 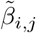 *which are usable only for population distribution of China, since we have used the data collected in China. But using the relationship* 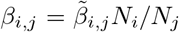, *as defined in Section A*.*1*.*4, we can obtain normalised values that can be used for all population densities, where N*_*i*_, *N*_*j*_ *are the population ratios in age-groups i and j. For each other target population we can use the equation* 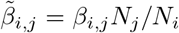 *to calculate the* 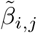 *in* (9) *and then solve the ODEs to obtain the evolution of the trajectories*.

**Note A.2.2** *This methodology can be applied to any uncontained or contained population if the effective basic reproduction number, R*_0_ *is known. But given the difficulty in estimating R*_0_ *in a contained population, the data collected in early stages of the spread of the virus in China seems to still be one of the best available options for the optimisation scheme*.

### A.3 Optimising the distribution of limited vaccine units

This Section briefly explains the optimisation scheme used in Section 3.4 to optimise the distribution of a limited number of vaccine units in the population. The optimisation algorithm used for this problem is the exact same global optimisation method used in Section A.2 and I will not repeat its details. But the objective function is obviously different. In this case, the objective function we aim to minimise is a weighted sum of two terms. The main term, is the final aggregate value of the Recovered compartment. As a reminder, the value of compartment *R* at each time represents the total number of those who were infected at any time in the past and are not Infectious any more. The second term is devised to make sure the total number of vaccine units remains constant. Simply, turning a constrained optimisation problem into an unconstrained optimisation problem to save computational time. The Matlab^*Q*c^ script used for this optimisation scheme can also be found in [9]. The optimisation algorithm runs in around 5-6 minutes on 40 hyper-threaded CPUs of type Intel(R) Xeon(R) CPU E5-2687W v4 @ 3.00GHz.

### A.4 SIR Case

As already mentioned in Section 1, the SEIR model is a suitable choice for the spread of SARS-CoV-2 in a population. But when I did my initial work on the spread of SARS-CoV-2 back in March 2020, very little was known about the dynamics of the virus and an SIR model seemed suitable. Nevertheless, as a thought experiment, in this section we can have a look at how the SARS-CoV-2 would spread in a population if instead of 4.6 days in Exposed period and 5 days in Infectious period, there is no Exposed period and there exists 9.6 days of Infectious period. Table 6 shows the eventual Recovered and maximum Infectious ratio of the same countries as in Table 1, but using an SIR model for the spread of SARS-CoV-2. Comparing the two tables we can see that the eventual Recovered ratio is less in SIR case and the peak of instantaneous Infectious has decreased.

**Table 6:** Maximum instantaneous Infectious ratio and eventual Recovered ratio in different countries in the uncontained scenario for SIR model.

| Country | Eventual Ratio of Recovered Population (%) | Maximum Instantaneous Infectious Ratio (%) |
| --- | --- | --- |
| Germany | 81.80% | 18.40% |
| Iran | 71.31% | 13.61% |
| Italy | 81.84% | 18.38% |
| Spain | 82.01% | 18.40% |
| China | 77.39% | 15.71% |
| USA | 81.20% | 17.87% |
| UK | 82.07% | 18.34% |
| France | 82.81% | 18.76% |
| South Africa | 69.20% | 12.80% |

Figures 5a and 5b shows the evolution of Infectious and Recovered ratios in these countries.

**Figure 5:**
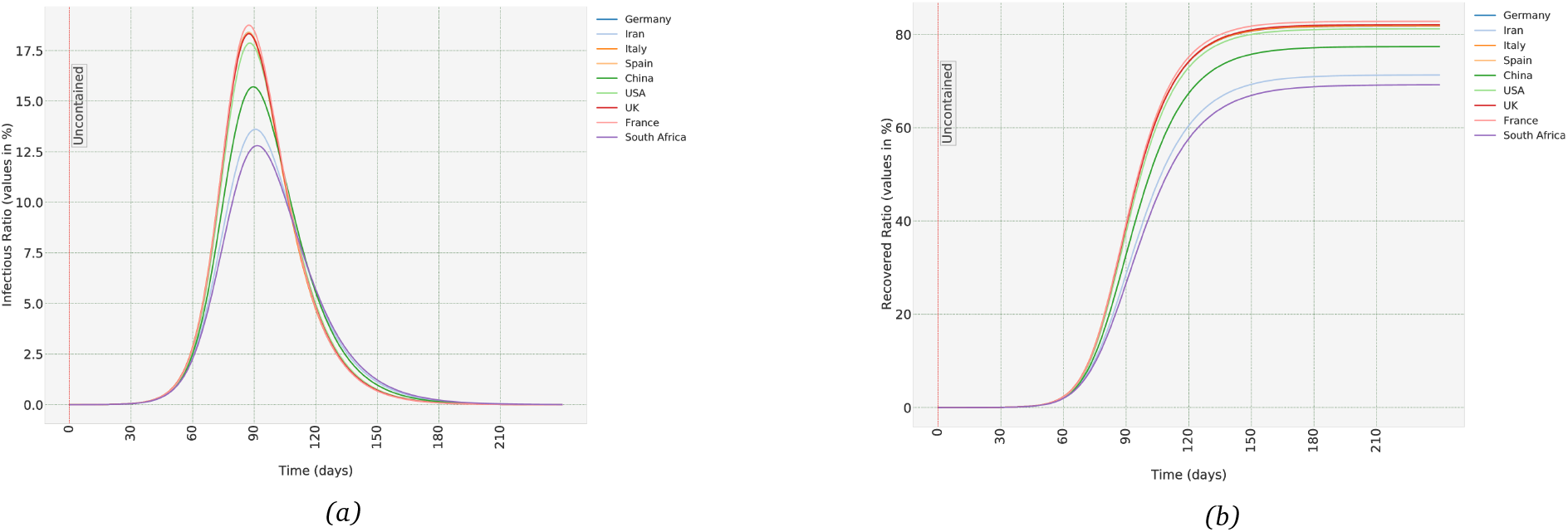
(a) The uncontained scenario in different countries assuming SARS-CoV-2 dynamics follow an SIR model, not an SEIR model, (a) Aggregate ratio of Infectious compartment, (b) Aggregate ratio of Recovered compartment.

### A.5 What was the Ratio of Asymptomatic Individuals Infected with SARS-CoV-2 in Diamond Princess Cruise-Ship?

A cruise ship named Diamond Princess was on the way to Yokohama in Japan and was supposed to dock on February 3rd 2020. On February 1st, a passenger who disembarked on January 25th to Hong Kong was tested positive for COVID-19. Subsequently, when the ship arrived in Yokohama on February 3rd, Japanese authorises forbade anybody from leaving the ship. Passengers were then informed that they should all go through a 14-day quarantine period between 5-19 February [24]. The virus spread through the ship and eventually, out of 3711 people onboard, 691 were tested positive [25], of which 13 lost their lives [26]. You can read the details of the events in [24] and [25], but what is of relevance in here is the disembarkation procedure. As explained in [25], a passenger was allowed to disembark the ship subject to:

- Completion of a 14-day period without sharing a cabin with a confirmed case; and
- Negative result for a SARS-CoV-2 by PCR in the final days of the period; and
- No relevant symptoms identified during a medical screening in the final day of the period.

This policy led to identification of many asymptotic cases in the ship, as summarised in Table 7. The age structure of people onboard, their mobility in the environment, and the manner of their interactions, even factors such as details of the ventilation system of the ship can affect who was infected and who was spared. But the ratio of infected people that were asymptomatic or symptomatic, to the best of my knowledge, is not affected by such environmental factors. That ratio depends on the dynamics of the virus and how it interacts with the host. With that in mind, it is not unreasonable to assume the values reported in Table 7 to be a rough estimate for the asymptomatic to total infected ratios for corresponding age-groups in any other human population.

**Table 7:** Data obtained from Diamond Princess cruise ship [25]. For age-groups 0-9 and 10-19, the total number of infected is too low, hence I have used the values obtained for age-group 20-29 as a substitute.

| Age Group | Confirmed Symptomatic Cases | Confirmed Asymptomatic Cases | Total Confirmed Cases | Personas Aboard as of Feb 5th | Ratio of Symptomatic to All | Ratio of Asymptomatic to All |
| --- | --- | --- | --- | --- | --- | --- |
| **0-9** | 0 | 1 | 1 | 16 | **89.29%**<br>(original 0.00%) | **10.71%**<br>(original 100.00%) |
| **10-19** | 2 | 3 | 5 | 23 | **89.29%**<br>(original 40.00%) | **10.71%**<br>(original 60.00%) |
| **20-29** | 25 | 3 | 28 | 347 | **89.29%** | **10.71%** |
| **30-39** | 27 | 7 | 34 | 428 | **79.41%** | **20.59%** |
| **40-49** | 19 | 8 | 27 | 334 | **70.37%** | **29.63%** |
| **50-59** | 28 | 31 | 59 | 398 | **47.46%** | **52.54%** |
| **60-69** | 76 | 101 | 177 | 923 | **42.94%** | **57.06%** |
| **70-79** | 95 | 139 | 234 | 1015 | **40.60%** | **59.40%** |
| **80+** | 29 | 25 | 54 | 227 | **53.70%** | **46.30%** |
| **Total** | 301 | 318 | 619 | 3711 | N/A | N/A |

A problem in using the age-stratified data recorded in Diamond Princess (Table 7) is that numbers of individuals onboard and infected for age-groups 0-9 and 10-19 are too low and they cannot be used as the basis for a reliable estimate. For those, we can use the ratio obtained for age-group 20-29 as a substitute. Having these values, we can then easily calculate the overall asymptomatic and symptomatic ratios in any population with a known age structure. For example, the overall asymptotic ratio for Germany and Iran, as representative of countries with relatively older and younger populations, respectively, can be calculated based on the population distribution of each. The result is that in Germany, 65.96% of infected cases would be symptomatic and 34.04% asymptomatic. In Iran, the ratios are 75.84% and 24.16%, respectively. That shows the extent to which this ratio changes based on the age structure of a population, and that countries with older populations have a higher asymptomatic ratio.

